# The Bern Sleep-Wake Registry: A 25-year multimodal resource for data-driven sleep medicine

**DOI:** 10.64898/2026.10.09.26364965

**Authors:** Julia van der Meer, Irina Filchenko, Anne-Kathrin Brill, Elena Wenz, Livia Fregolente, Kseniia Zub, Anelia Dietmann, Carolin Schäfer, Sandra C. Röthlisberger, Annina Helmy, Athina Tzovara, Luigi Fiorillo, Francesca Faraci, Waldo Valenzuela, Kaspar Schindler, Wolfgang Schmitt, Markus H. Schmidt, Claudio L.A. Bassetti

## Abstract

**Background:** Sleep–wake-circadian disorders are prevalent and contribute substantially to morbidity, mortality and reduced quality of life. Evaluation in real-world settings is essential to capture their complexity and association with long-term outcomes. Most sleep-wake registries are restricted to selected populations, lack systematic integration of objective sleep-wake-circadian assessments with the clinical data and provide limited follow-up. To address these limitations, we established the Bern Sleep-Wake Registry based on real-world data from a tertiary University Hospital sleep center.

**Methods:** The registry includes data from patients referred to the multidisciplinary Sleep-Center at Inselspital, University Hospital Bern (founded in 1990), from the year 2000 onwards who underwent at least one objective assessment such as polysomnography, daytime sleep-wake recordings, vigilance tests, and actigraphy. The registry integrates clinical information, questionnaire scores, outputs of objective assessments and raw signal data, and is regularly updated to incorporate new patients and longitudinal follow-up information.

**Results:** As of September 2025, records from over 11’000 patients were evaluated (median age: 50 years (interquartile range 36, 61); 65% male). The most common sleep-wake-circadian disorders were “sleep-related breathing disorders” (67%), “insomnia disorders” (15%) and “central disorders of hypersomnolence” (9%). Available assessments included more than 12’000 polysomnographies, 2’400 multiple sleep latency tests, 2’100 maintenance of wakefulness tests, 1’800 vigilance tests and 4’500 actigraphies.

**Conclusions:** By longitudinally integrating multimodal deep clinical phenotyping with objective assessments and raw physiological signals, the Bern Sleep-Wake Registry provides a foundation for investigating disease trajectories, identifying novel biomarkers and developing data-driven approaches to precise diagnosis, prognosis and individualized treatment.

## 1 Introduction

Sleep is a fundamental physiological process integral to body and mind. Healthy sleep supports memory consolidation, emotional regulation, immune functions, autonomic balance and vascular function^1,2^. Sleep-wake-circadian disorders (SWCD) are highly prevalent, affecting chronically more than 30% of the general population worldwide^3^. They contribute to increased morbidity as well as reduced quality of life and are increasingly recognized as risk factors for adverse cognitive outcomes, vascular events as well as for endocrine, metabolic, autoimmune, and neoplastic diseases^4–6^.

Given that the relationships between sleep and physiological functions span across multiple organ systems and evolve over time, evaluation of real-world large-scale data is essential to capture the complexity of SWCD burden and its association with long-term outcomes. In sleep medicine, many promising interventions failed, often due to underappreciated variability among individuals, limited generalizability, or unrecognized phenotypes^7,8^. Real-world data offer insights into the heterogeneity, comorbidities, and trajectories of disease that are often missed in highly controlled trials, whereas large-scale data enhances equity and generalizability through the detection of rare subgroups and subtle patterns.

In specialized sleep clinics, multimodal assessments are carried out, but associated data are not usually accessible for research due to technical and data privacy constraints. Over the past years, efforts have been made to establish sleep-oriented registries^9–21^. However, existing sleep-oriented registries have important limitations. Many are restricted to circumscribed populations (e.g., pediatric or aging cohorts) or focus on specific disorders (e.g., obstructive sleep apnea). Few integrate raw signal data from objective sleep-wake-circadian assessments (e.g., polysomnography (PSG), actigraphy, or vigilance tests) jointly with detailed clinical and questionnaire data, and most lack SWCD diagnoses and longitudinal follow-up. An overview on the most relevant large-scale sleep registries is depicted in **Table 1**. To address these gaps, we established the Bern Sleep-Wake Registry (BSWR) – a real-world large-scale registry embedded in a tertiary, multidisciplinary sleep center which was launched in 1990 at Inselspital, University Hospital of Bern, Switzerland. The aim of the BSWR is to provide an interoperable repository of multimodal data, which is easily accessible for algorithmic exploitation, updatable to support follow-ups extending over decades as well as growth of the cohort over time, and compliant with standards for data privacy. The BSWR integrates longitudinal objective sleep-wake-circadian assessments, clinical data, living status, standardized questionnaires, and laboratory results.

**TABLE 1.** Characteristics of selected sleep-wake-circadian datasets. Selected databases available via the National Sleep Research Resource are indicated as separate entries, with indication of accessibility via NSSR.

| Name | Location | Sample | Population | PSG | Actigraphy | MSLT | MWT | Questionnaires | Lab | RPG | Accessibility |
| --- | --- | --- | --- | --- | --- | --- | --- | --- | --- | --- | --- |
| National Sleep Research Resource<sup>9</sup> | USA (Multi-Study) | ~26,808 | Highly diverse (many cohorts), Ages 4–97 | Yes | Yes | Ø | Ø | Yes | Yes | Yes | Open |
| Sleep Heart Health Study<sup>10</sup> | USA (Multi-center) | 6'441 | Ages >= 40 | Yes | Ø | Ø | Ø | Yes | Yes | Ø | via NSSR |
| Nationwide Children's Hospital DataBank<sup>14</sup> | USA | 3'673 | Pediatric | Yes | Ø | Ø | Ø | Ø | Ø | Ø | via NSSR |
| Multi-Ethnic Study of Atherosclerosis<sup>15</sup> | USA (Multi-center) | 2'230 | Ages 54-93 | Yes | Ø | Ø | Ø | Yes | Ø | Ø | via NSSR |
| Wisconsin Cohort Study<sup>16</sup> | USA | 1'500 | State employees | Yes | Yes | Yes | Ø | Yes | Yes | Ø | via NSSR |
| UK Biobank<sup>17</sup> | United Kingdom (Multi-center) | ~90'000 | Ages 40-69 | Ø | Yes | Ø | Ø | Yes | Yes | Ø | On request |
| HypnoLaus<sup>18</sup> | Switzerland | 1'172 | Ages 40-85 | Yes | Ø | Ø | Ø | Yes | Ø | Ø | On request |
| European Sleep Apnea Database<sup>19</sup> | Europe (33 countries) | >25,000 | Patients with OSA | Yes | Ø | Ø | Ø | Yes | Yes | Yes | Private |
| Chinese Clinical Sleep Database<sup>20</sup> | China (Multi-center) | 1'183 | Patients with SWCD | Yes | Ø | Yes | Ø | Yes | Yes | Ø | Private |
| Shanghai Sleep Health Study<sup>21</sup> | China | 5'748 OSA patients | Patients with OSA | Yes | Ø | Ø | Ø | Ø | Yes | Ø | Private |
| Japanese Database<sup>11</sup> | Japan (Multi-center) | 26'300 | Patients with SWCD | Yes | Ø | Ø | Ø | Ø | Ø | Ø | Private |
| Sydney Sleep Biobank<sup>12</sup> | Australia | 821 | Patients with SWCD | Yes | Ø | Ø | Ø | Yes | Yes | Ø | Private |

Abbreviations: Ø – not stated. PSG – polysomnography, MSLT – multiple sleep latency test, MWT – maintenance of wakefulness test, Lab – laboratory parameters, RPG – respiratory polygraphy, SDB – sleep disordered breathing. SWCD – sleep-wake-circadian disorders.

## 2 Methods

### 2.1 Setting: Sleep-Wake-Epilepsy Center of Inselspital, University Hospital of Bern

Following first polysomnographic recordings in 1982, the Sleep (later renamed Sleep-Wake-Epilepsy Center (SWEZ)) of the Inselspital, University Hospital Bern, was established officially as first multidisciplinary sleep center of Switzerland in 1990. Over the years, it developed into an internationally significant tertiary center for patient care, sleep research, and education, with a strong interdisciplinary approach. Based on their needs, SWCD patients consult with teams of neurologists, pulmonologists, psychiatrists, and pediatricians. Annually approximately 900 in-hospital PSGs are performed across four beds, alongside ~800 respiratory polygraphies. In addition, ~300 multiple sleep latency tests (MSLT), ~200 maintenance of wakefulness tests (MWT) with vigilance assessments, and ~500 actigraphies are conducted. Laboratory analyses (e.g., cerebrospinal fluid (CSF) hypocretin measurements, saliva melatonin, human leukocyte antigen (HLA) typing) complement the diagnostic spectrum. In 2024, SWEZ was officially recognized as reference center for rare SWCD by the Swiss authorities (coordination of rare diseases, Kosek^22^), within the framework of the Swiss Narcolepsy Network (SNaNe)^23^.

At the interface of clinical research and technological innovation, SWEZ maintains close collaborations with the Biobank of Inselspital Bern, the interdisciplinary research platform NeuroTec^24^, the ARTORG Center for Biomedical Engineering Research and the Institute of Computer Science at the University of Bern, the Biomedical Signal Processing group at the University of Applied Sciences and Arts of Southern Switzerland (SUPSI) and the Swiss Center for Electronics and Microtechnology (CSEM), all of which provide computational expertise to analyze the recorded data. SWEZ is also an important training site, providing education for medical students at the University of Bern, PhD candidates, and the International Master of Advanced Studies in Sleep Medicine^25^. Today, SWEZ is nationally and internationally recognized for its expertise in a broad spectrum of SWCD and continuously evolves to address emerging societal needs and support new technologies.

### 2.2 Study population

The here reported BSWR comprises patients referred to SWEZ between January 2000 and September 2025. While most of the patients reside in Canton of Bern, Switzerland, some patients come from other cantons or from abroad.

The BSWR was approved by the local ethics committee (Kantonale Ethikkommission Bern [KEK-Nr. 2022-00415]), ensuring compliance with the Human Research Act (HRA) and Ordinance on Human Research with the Exception of Clinical Trials (HRO). The BSWR ethics protocol builds on written patient consent for further use of their clinical data (general consent) and reflects a rolling-retrospective design covering usage of data for research purposes between 2000 and 2040. For research purposes, data are coded (i.e., all identifying information is removed) and filtered for patient consent according to the BSWR protocol. Only patients with at least one objective sleep-wake-circadian assessment (i.e., PSG, actigraphy, MSLT, MWT or vigilance task) are included. No further inclusion or exclusion criteria are applied. Data can be shared with external research groups upon reasonable request, involving a data transfer agreement describing terms of data usage and project scope.

### 2.3 History of the BSWR infrastructure

In 1982 first polysomnographic recordings were made at Inselspital in the Department of Neurology of the University of Bern. In 1990 the inter-multidisciplinary Sleep Center was officially inaugurated with one of the authors of this paper (CB) being among the first co-chairs. In 1993, the first version of the Bern sleep questionnaire and a first patient registry was introduced by one of the authors (CB) providing a systematic base for clinical assessments. The history of the Sleep Center is visualized in **Figure 1**.

**FIGURE 1.**
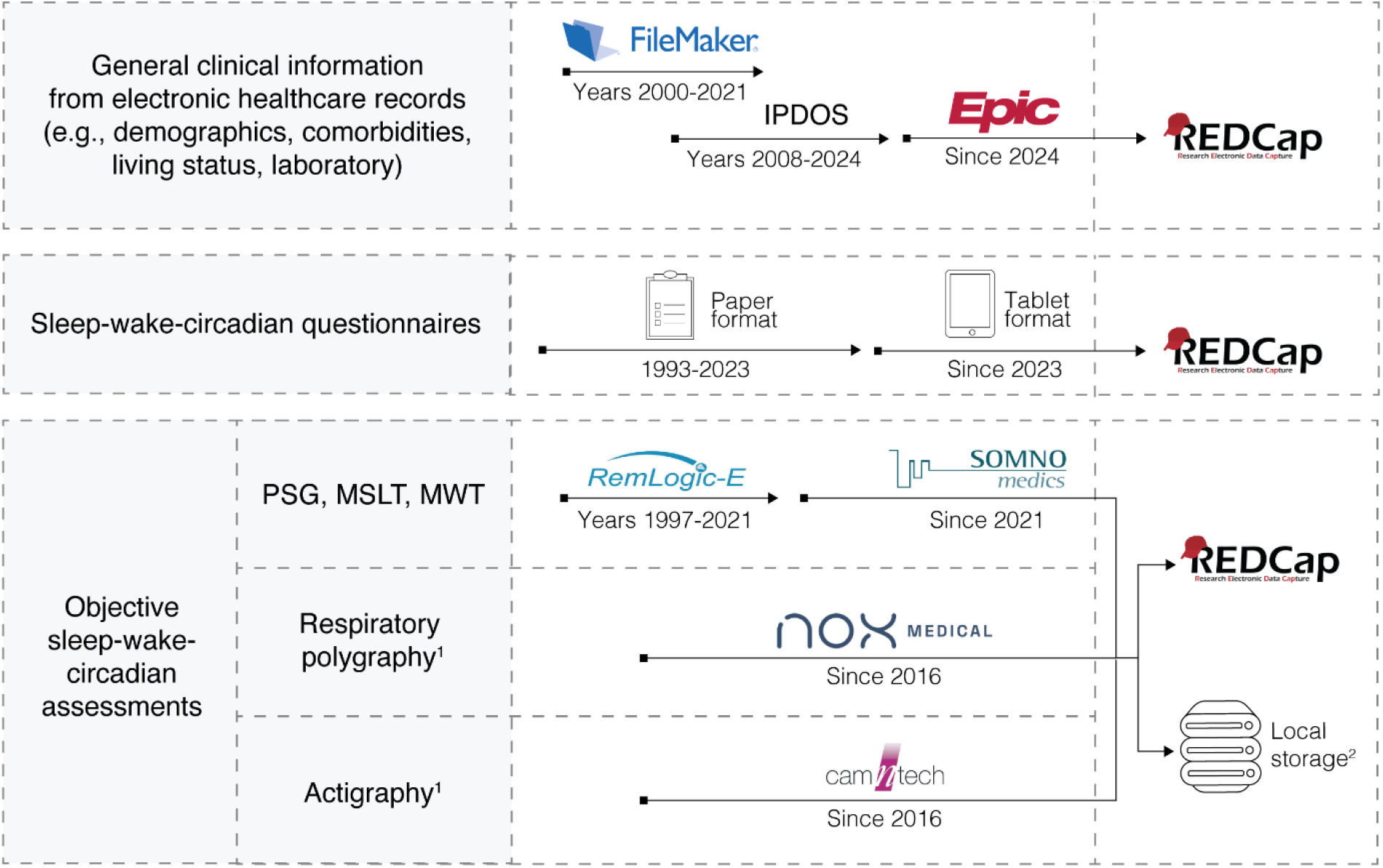
History of the BSWR registry. Graphical overview of the multimodal data collection including the three main modalities “general clinical information”, “sleep-wake-circadian questionnaires” and “objective sleep-wake-circadian assessments”. *Abbreviations: PSG – polysomnography, MSLT – multiple sleep latency test, MWT – maintenance of wakefulness test. Structured data are consolidated over time and modalities in a REDCap database*. ^*1*^*Data integration of Respiratory polygraphy and actigraphy planned in analogy to PSG data*. ^*2*^*Raw data are stored on a local storage. PSG raw data are further integrated in the Medical-Blocks platform*.

In 2000 a systematic multimodal collection of patient data at SWEZ started, using a FileMaker® clinical database, with PSG results transferred semi-automatically from the PSG recording system REMLogic™ (Embla Systems, Kanata, Canada) and all other multimodal information entered manually.

In 2016 historic data were consolidated in a REDCap database^26^ at the Clinical Trial Unit (CTU) Bern, and diagnoses for all records were manually re-classified to the American Academy of sleep medicine (AASM) International Classification of Sleep Disorders – Third Edition (ICSD-3)^27^ by medical experts. In the same year, MotionWatch actigraphy (CamNtech, Cambridgeshire, UK) and Noxturnal respiratory polygraphy (Nox Medical, Reykjavik, Iceland) were introduced as standard systems.

In 2020, the BSWR was expanded within the SPAS (Sleep Physician Assistant System) Eurostars-Horizon 2020 project with the aim to facilitate processing of sleep-related data in the clinical routine. This enabled systematic access to PSG raw data. In 2021, the PSG diagnostic system REMLogic™ was replaced by a SOMNO HD system (SOMNOmedics AG, Randersacker, Germany) and automated retrieval of clinical information from the electronic health care systems (EHCS) was implemented in collaboration with the Insel Data Science Center (IDSC). With this, an HL7-based data exchange between clinical and PSG systems and a python-based pipeline for the integration of historic and newly introduced data sources into a unified REDCap database were established.

Since 2022, clinical data, PSG raw data, and associated algorithms have been maintained on the *Medical-Blocks-A Platform*^28^ at the Neurocenter of Inselspital. Medical-Blocks is a research data platform developed at Inselspital for the secure management, de-identification, and analysis of multimodal clinical and imaging data, providing role-based access control, audit logging, and standardized export formats. It is operated as two synchronized instances: MB-Insel, hosted within the Inselspital network and connected to the hospital clinical systems, and MB-UniBe, hosted at the University of Bern for collaboration with external partners across the different international projects of the sleep clinic in line with FAIR principles^29^. We plan to convert respiratory polygraphy and actigraphy raw data to de-identified standard formats and store them on the *Medical-Blocks-A Platform* in analogy to the PSG raw data. To further enhance the multimodal BSWR dataset in future projects, existing MRI data from SWCD patients can be made available via the Medical-Blocks-A Platform.

The “Bern Sleep Health Questionnaire” was introduced in 2020, and since 2023 patients complete questionnaires in the waiting room before their clinical visit via REDCap surveys providing automated summary reports. In 2024, with the roll-out of EPIC, the hospital-wide EHCS at Inselspital, the SOMNOmedics HL7 API was further expanded to support bidirectional transfer of sleep study orders and structured PSG results.

### 2.4 Major data types and clinical workflow

The major data types include demographics, medical history, living status, questionnaires, objective sleep-wake-circadian assessments (i.e., respiratory polygraphy, PSG, MSLT, MWT, vigilance test, actigraphy), laboratory values, and SWCD diagnoses (**Figure 2**). Patients referred to the SWEZ usually undergo a clinical workflow, examples are illustrated in **Figure 3**.

**FIGURE 2.**
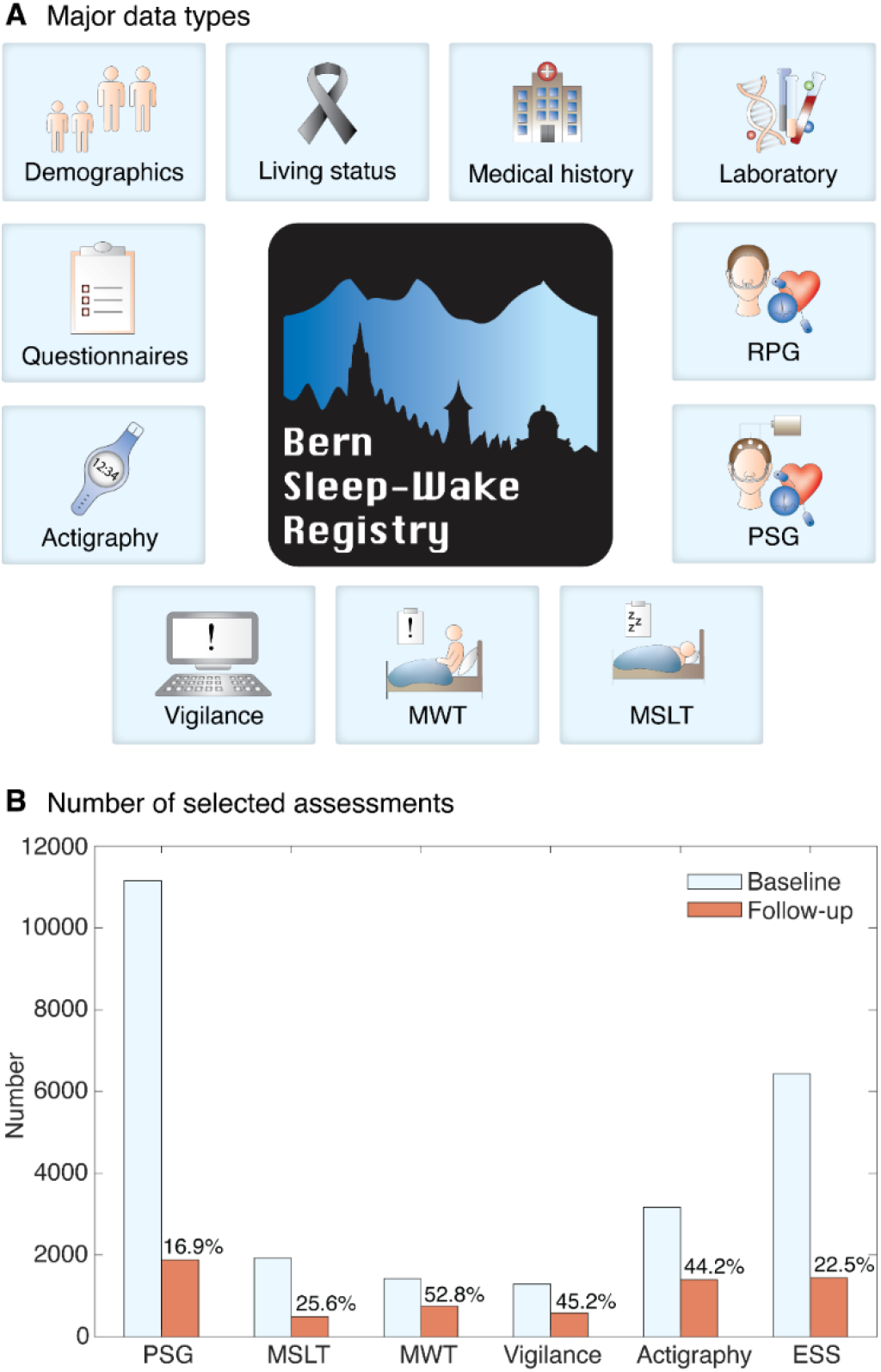
**A**. Major data types in the Bern Sleep-Wake Registry. **B**. Number of selected assessments at baseline and follow-up visits (as of September 2025). *Abbreviations: PSG – polysomnography, MSLT – multiple sleep latency test, MWT – maintenance of wakefulness test, RPG – respiratory polygraphy, ESS – Epworth Sleepiness Scale*.

**FIGURE 3.**
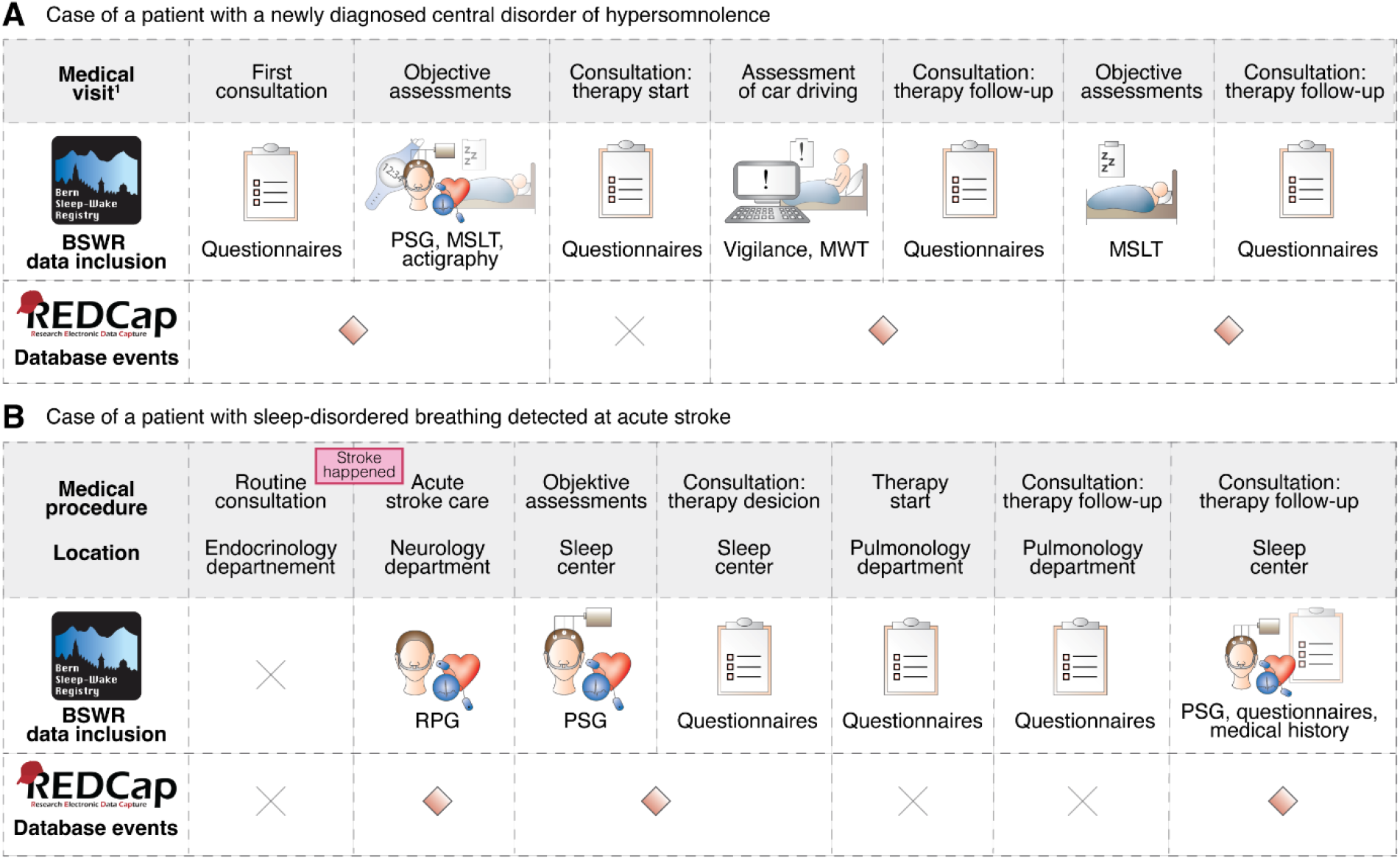
**A**. Case of a patient with a central disorder of hypersomnolence. At the first consultation, the patient completes questionnaires to document initial symptoms. Objective sleep-wake-circadian assessments, which may include PSG, MSLT and actigraphy, are then scheduled in the nearest time (i.e., the first event in the REDCap database). In the weeks following the assessment, a consultation is held to establish a diagnosis and start therapy. The patient typically completes follow-up sleep questionnaires to monitor symptoms. Depending on individual needs, an assessment of driving ability may be scheduled, usually followed by another consultation to discuss therapy and track symptoms (i.e., the second event in the REDCap database). Over time, additional objective sleep assessments (e.g., MSLT) and consultations may be performed, for example after one year, to document symptom progression (i.e., the third or later event in the REDCap database). **B**. Case of a patient with sleep-disordered breathing detected at acute stroke. The patient is referred emergently to stroke care, where an RPG is performed during acute stroke to screen for SDB (i.e., first REDCap event). If moderate to severe SDB is detected on RPG, the patient is referred to the sleep center for reevaluation of the situation, potentially indicating further diagnostics including PSG, the gold standard for diagnosing sleep-disordered breathing. After PSG, the patient attends a consultation at the interdisciplinary sleep center to discuss therapy (i.e., second REDCap event). The patient may then be referred to pulmonology, where questionnaires may be collected again at therapy initiation and follow-up, but these are typically not entered in REDCap since no objective sleep assessment is performed during this period. Finally, the patient may return to the sleep center for follow-up PSG, for example to titrate positive airway pressure therapy, where questionnaires and clinical assessments are completed again (i.e., the third REDCap event). ^1^Location for all medical visits: sleep center. *Abbreviations: PSG – polysomnography, MSLT – multiple sleep latency test, MWT – maintenance of wakefulness test, RPG – respiratory polygraphy, SDB – sleep-disordered breathing*.

#### 2.4.1 | Demographics, medical history, living status

Global patient information including demographics, medical history, and living status are obtained from the EHCS via the IDSC. Parameters are listed in **Table S1**, standardized questionnaires are listed in **Table S2**.

Medical history provides essential information for clinical research, typically including details on comorbidities, medications, and treatments. Currently, our data management efforts focus on retrieving structured information on comorbidities and medication from EPIC. Complementary, we use natural language processing to extract ICD-11 clinical diagnoses and onset dates for major diseases of interest (e.g., stroke) from textural reports of historical records, with expected completion in 2026^30^. We plan to retrieve additional structured and free text clinical information from EPIC, including therapies and treatment success. For the long-term, we aim to enable longitudinal assessment of health trajectories by establishing a standardized data model, based on existing common data elements, to improve interoperability of sleep-wake-circadian related data^31^.

#### 2.4.2 Objective sleep-wake-circadian assessments

Objective sleep-wake-circadian assessments include respiratory polygraphy, PSG, MSLT, MWT, vigilance test, and actigraphy.

To date, our efforts focused on PSG, which is performed for most patients at SWEZ. Depending on the clinical indication, patients also undergo MSLT and MWT, with intermittent vigilance tests following the overnight PSG with set-up and montage according to AASM recommendations. Initial scoring according to AASM guidelines^32^ is performed by technicians and then reviewed by qualified medical doctors, which are typically certified somnologists. We further plan to integrate automatic sleep scoring and quantification of sleep microarchitecture to the clinical routine^33,34^. Parameters from overnight PSG and daytime MSLT and MWT are extracted from the SOMNOmedics system. Overnight PSG parameters are listed in **Table S3**. Parameters from the assessments of daytime sleepiness are detailed in **Table S4**.

Respiratory polygraphy analysis is performed in Noxturnal, and the automated scoring is manually corrected by qualified medical doctors according to AASM guidelines. Actigraphy is usually conducted for two to three weeks before the PSG night using MotionWatch software, which records sleep diaries, light exposure, and activity. Actigraphy parameters, including results from non-parametric circadian rhythm analysis, are provided by the MotionWatch software package^35^ and manually entered to EPIC. As a next step, we plan a pipeline for the transfer of these actigraphy results to the REDCap database. Actigraphy parameters are listed in **Table S5**. Parameters on general information from sleep-wake-circadian assessments are listed in **Table S6**.

#### 2.4.3 Laboratory values

Currently, lab-derived parameters are available only for selected patient groups (e.g. HLA type and orexin levels in suspected narcolepsy). We plan to also transfer laboratory values from the EHCS into REDCap, including basic clinical parameters (e.g. blood count, cholesterol, HbA1c) and SWCD-specific parameters (e.g. orexin, ferritin, iron).

#### 2.4.4 Sleep-wake-circadian disorder classification

As the ICSD-3 has been criticized for insufficiently integrating biomarkers, particularly for central disorders of hypersomnolence^36^, we report three additional categories: “Excessive daytime sleepiness not otherwise specified” for evidence of a central disorder of hypersomnolence not meeting criteria for any subtype (e.g. narcolepsy type 2, idiopathic hypersomnia); “Poor sleep hygiene” for inconsistent sleep patterns not meeting criteria for a circadian rhythm sleep-wake disorder (e.g. shift work disorder, jet lag disorder); and “Periodic limb movements of sleep” if explicitly mentioned in the clinical diagnosis without meeting criteria for a sleep-related movement disorder (e.g. periodic limb movement disorder, restless legs syndrome). Together with the ICSD-3 sleep disorders, these are summarized as “Reported sleep problems”. SWCD-associated parameters are listed in **Table S7**.

At the baseline visit defined as the first examination in our sleep lab, median age was 50 (36, 61) years. Most patients were adult, with the “middle adulthood” group (40–64 years) comprising 53% of the study population (**Figure 4A**). Nearly two third of the patients were male (65%), reflecting the marked overrepresentation of men starting from “middle adulthood” (**Figure 4B**).

**FIGURE 4.**
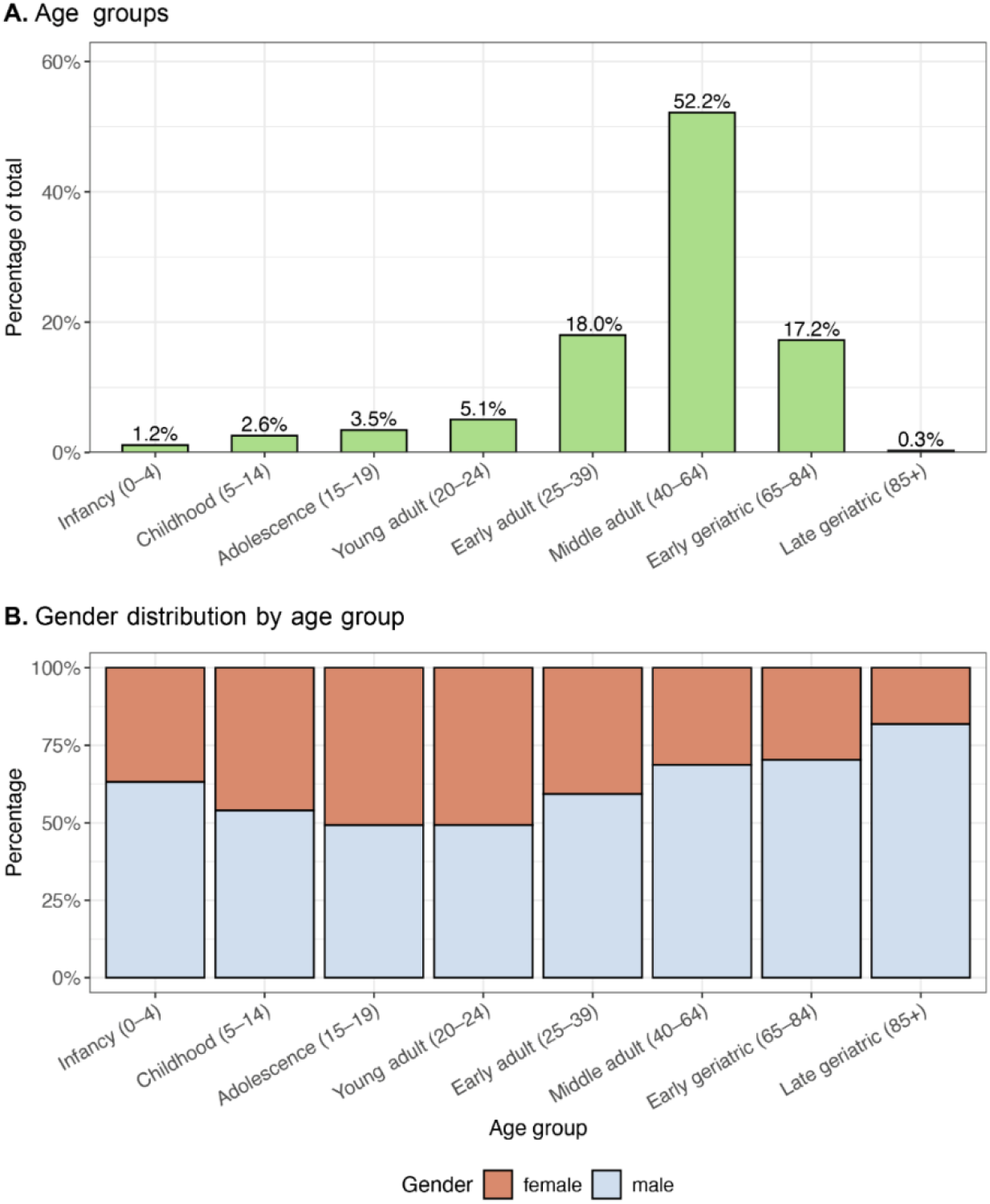
A. Age groups according to International Classification of Diseases 11th Revision (ICD-11) classification. B. Gender distribution by age groups.

### 2.5 Data analysis

Given the primary aim of describing the database, analyses were limited to descriptive statistics. Categorical variables were expressed as frequencies and percentages, continuous variables as medians with interquartile ranges (IQRs). More detailed, stratified analyses by age, sex, and disorder will be reported in a subsequent publication. All analyses were conducted using R version 4.4.2.

## 3 Results

### 3.1 Demography

Until September 30, 2025, data of totally 11’656 patients with 14’786 visits from the BSWR were included for scientific evaluation according to general consent based ethical approval. Overall and gender stratified characteristics of the study population are shown in **Table 2**.

**TABLE 2.** Selected characteristics of BSWR population (September 2025).

| Characteristic | Overall |  | Gender stratified |  |
| --- | --- | --- | --- | --- |
|  | N | N = 11,656<sup>1</sup> | male<br>N = 7,528<sup>2</sup> | female<br>N = 4,128<sup>2</sup> |
| Gender (Male) | 11,656 | 7,528 (65%) |  |  |
| Age (Years) | 11,656 | 50 (36, 61) | 52 (39, 62) | 48 (31, 59) |
| BMI (kg/m<sup>2</sup>) | 6,716 | 26 (23, 30) | 27 (24, 30) | 25 (22, 30) |
| ESS | 6,574 | 10 (6, 14) | 9 (5, 13) | 11 (6, 14) |
| FSS | 2,055 | 4.70 (3.30, 5.70) | 4.30 (3.00, 5.30) | 5.00 (3.90, 6.00) |
| PSG sleep duration (minutes) | 11,156 | 344 (296, 389) | 343 (295, 387) | 346 (300, 391) |
| PSG sleep efficiency (%) | 11,106 | 87 (77, 94) | 87 (77, 94) | 88 (78, 94) |
| PSG sleep latency (minutes) | 11,130 | 11 (5, 22) | 10 (5, 20) | 13 (6, 26) |
| PSG NREM1 (%) | 11,010 | 17 (11, 27) | 19 (12, 29) | 14 (9, 21) |
| PSG NREM2 (%) | 11,011 | 47 (39, 55) | 48 (39, 56) | 47 (39, 54) |
| PSG NREM3 (%) | 11,011 | 17 (9, 25) | 15 (7, 23) | 21 (14, 29) |
| PSG REM (%) | 10,998 | 15 (10, 20) | 15 (10, 19) | 15 (10, 20) |
| PSG AHI (events/h sleep) | 11,108 | 12 (4, 28) | 16 (7, 34) | 7 (2, 17) |
| PSG PLMS Index (events/h sleep) | 9,099 | 3 (0, 14) | 3 (0, 16) | 2 (0, 9) |
| MSLT sleep latency (minutes) | 1,932 | 7 (4, 11) | 7 (4, 11) | 7 (4, 11) |
| MWT sleep latency (minutes) | 1,423 | 32 (17, 40) | 34 (19, 40) | 29 (16, 40) |
| PVT mean RT (ms) | 1,290 | 279 (243, 348) | 268 (240, 324) | 294 (253, 372) |
| Actigraphy Inactivity Index (%) | 3,157 | 35 (31, 40) | 34 (30, 40) | 37 (33, 41) |

<sup>1</sup>n (%); Median (Q1, Q3)
<sup>2</sup>Median (Q1, Q3)
Abbreviations: BMI – body mass index, ESS – Epworth sleepiness scale, FSS – Fatigue severity scale, PSG – polysomnography, REM – rapid eye movement sleep, AHI – apnea hypopnea index, PLMS – periodic limb movements during sleep, MSLT – multiple sleep latency test, MWT – maintenance of wakefulness test, PVT mean RT – Psychomotor vigilance test mean reaction time.

### 3.2 Multimodal sleep-wake-circadian assessments

At the baseline visit, BMI was 26 kg/m^2^ with a prevalence of obesity (BMI>=30 kg/m^2^) of 25%. ESS and FSS scores were 10 and 4,7 respectively. PSG derived sleep duration was 344 minutes with a sleep latency of 11 minutes and a sleep efficiency of 87%. AHI was 12/h with a prevalence of 44% for AHI>15/h and 23% AHI>30/h, respectively. PLMI was 3/h with a prevalence of 23% for PLMI>15/h. MSLT and MWT sleep latencies were 7 and 32 minutes, respectively. Psychomotor vigilance test (PVT) mean reaction time was 279 ms. Actigraphy inactivity index was 35%. Overall and gender stratified values are listed in **Table 2**.

### 3.3 Diagnoses

The prevalences of ICSD-3 SWCD categories in the BSWR cohort (n=11’656) were: sleep-related breathing disorders (67%), insomnia disorders (15%), central disorders of hypersomnolence (9.1%), sleep-related movement disorders (8.1%), parasomnias (4.6%) and circadian rhythm sleep-wake disorders (1.2%). Clinically relevant “poor sleep hygiene” and “periodic limb movements of sleep” as defined in section 2.4.4 were frequent (each 14%). Overall, 85% of the patients were assigned to at least one SWCD according to ICSD-3, and SWCD comorbidities were frequent, with 19% of the patients having been assigned more than one ICSD-3 diagnosis. Overall and gender stratified values are listed in **Table 3**.

**TABLE 3.** The prevalence of sleep-wake-circadian disorders in the BSWR population (September 2025).

| Characteristic | Overall | Gender stratified |  | p-value<sup>2</sup> |
| --- | --- | --- | --- | --- |
|  | N = 11,656<sup>1</sup> | male<br>N = 7,528<sup>1</sup> | female<br>N = 4,128<sup>1</sup> |  |
| Insomnia disorders | 1,798 (15%) | 1,000 (13%) | 798 (19%) | <0.001 |
| Sleep-related breathing disorders | 7,838 (67%) | 5,691 (76%) | 2,147 (52%) | <0.001 |
| Central disorders of hypersomnolence | 1,060 (9.1%) | 566 (7.5%) | 494 (12%) | <0.001 |
| Circadian rhythm sleep-wake disorders | 140 (1.2%) | 90 (1.2%) | 50 (1.2%) | >0.9 |
| Parasomnias | 535 (4.6%) | 346 (4.6%) | 189 (4.6%) | >0.9 |
| Sleep-related movement disorders | 939 (8.1%) | 573 (7.6%) | 366 (8.9%) | 0.017 |
| Excessive daytime sleepiness not otherwise specified | 598 (5.1%) | 261 (3.5%) | 337 (8.2%) | <0.001 |
| Poor sleep hygiene | 1,596 (14%) | 920 (12%) | 676 (16%) | <0.001 |
| Periodic limb movements of sleep | 1,654 (14%) | 1,238 (16%) | 416 (10%) | <0.001 |
| ICSD-3 Sleep disorders | 9,863 (85%) | 6,673 (89%) | 3,190 (77%) | <0.001 |
| ICSD-3 Sleep disorders (N>1) | 2,186 (19%) | 1,421 (19%) | 765 (19%) | 0.6 |
| Reported sleep problems | 10,698 (92%) | 7,048 (94%) | 3,650 (88%) | <0.001 |
| Reported sleep problems (N>1) | 4,549 (39%) | 2,998 (40%) | 1,551 (38%) | 0.017 |

<sup>1</sup>n (%) <sup>2</sup>Pearson's Chi-squared test
Abbreviations: ICSD-3 – International Classification of Sleep Disorders - Third Edition.

## 4 Discussion

The BSWR is an ongoing registry representing the environment of a specialized tertiary sleep center with expertise in neurology, psychiatry, and pneumology. It ensures accessibility of multimodal sleep-wake-circadian related data for both clinical (i.e., patient-centric) and research applications, scalability, updatability as well as technical and ethical frameworks for data privacy and security. The BSWR provides an environment for the development and validation of new technologies and could be a model for other sleep centers, since it presents solutions for common challenges in clinical and research data management.

By encompassing patients with a broad range of SWCD and comorbidities, the BSWR constitutes a highly heterogeneous dataset reflecting real-world practice. In contrast to most existing registries, which typically provide structured summaries or selected outcome measures (**Table 1**), the BSWR is among the few repositories offering large-scale access to objective sleep assessments combined with detailed clinical records, including long-term follow-up. In Switzerland, also the HypnoLaus database provides large-scale, multimodal sleep-health data with longitudinal follow-up^18^. Compared to HypnoLaus, which focuses on the general population aged 40 to 85 years, BSWR captures a patient population assessed in a specialized tertiary sleep center. Together, these registries provide complementary patient- and population-based samples from the Swiss population, supporting a broad range of sleep research applications.

In the BSWR, the high proportion of patients aged 40–64 years may reflect the cumulative impact of occupational, social, and medical burden in midlife, as well as center specific referral patterns. This life period is characterized by high cumulative stress, body weight gain and increasing cardiometabolic comorbidities, all of which might contribute to the high prevalence of SDB mostly attributable to middle-aged men^16^. In contrast, female-dominated disorders like insomnia might be underrepresented in our database because PSG and other objective SWCD examinations are not refunded by health insurances for patients with insomnia in Switzerland. In the BSWR, overlapping classifications of ICSD-3 SWCD categories occur in ~20% of the patients. This emphasizes the real-world complexity of SWCD, the clinical importance of a holistic view and individualized, multimodal phenotyping, as multiple coexisting SWCD and other morbidities may interact to worsen symptom burden and affect treatment response. While the frequencies reported here are consistent with our clinical experience, comparison with earlier reports is hampered by changes in SWCD classification, conceptual shifts like reporting prevalence of disorders versus primary disorders, cultural effects on sleep disorders and their diagnosis, as well as center-specific referral biases.

With the aim to support applications spanning from diagnosis support and long-term prognosis to knowledge dissemination and technological transformation, the BSWR is continuously adapting to the challenges of clinical care (**Figure 5**).

**FIGURE 5.**
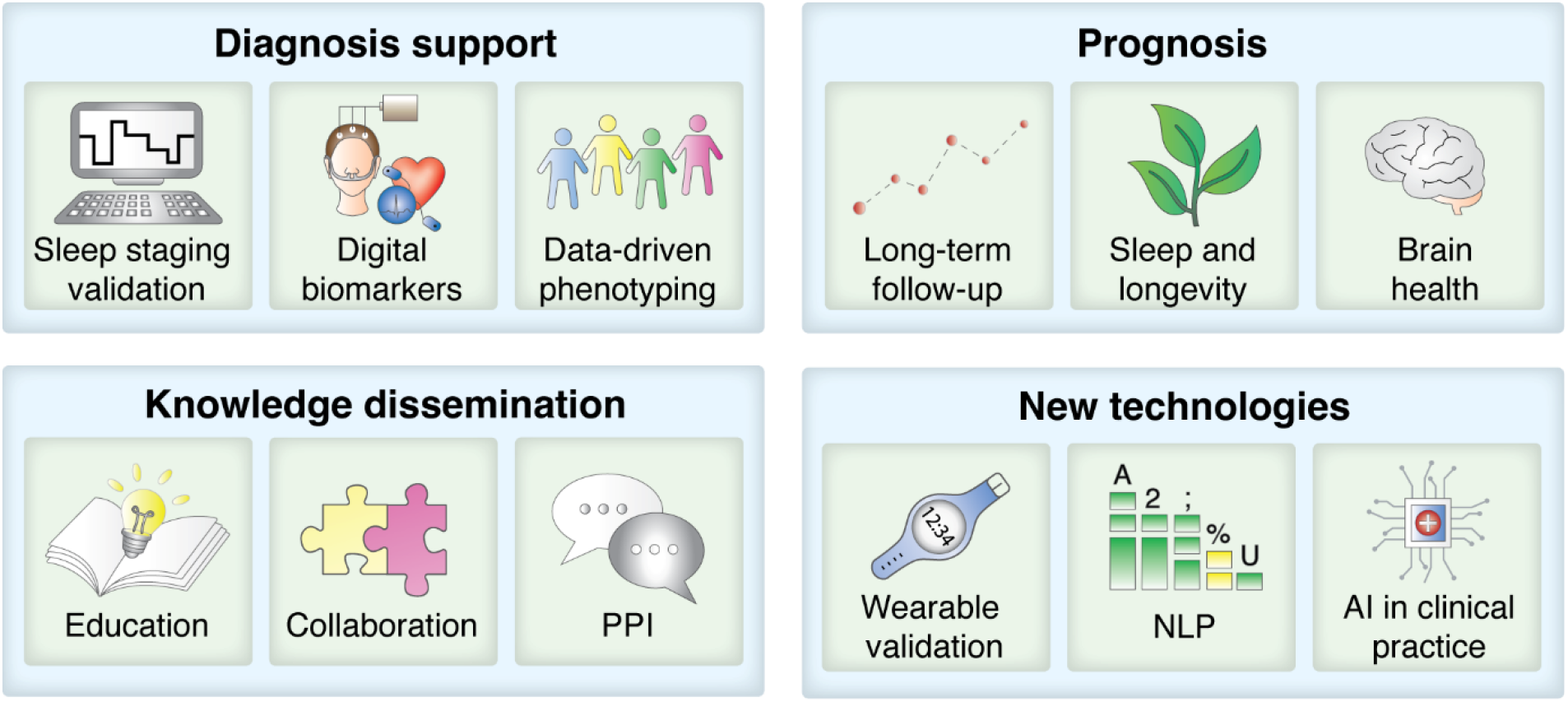
BSWR Applications. The BSWR facilitates validation of sleep scoring methodologies, identification of digital biomarkers, and data-driven phenotyping to support diagnosis. Longitudinal patient follow-up enables the assessment of long-term disease trajectories and prognosis. In addition, the BSWR serves as a central hub for knowledge dissemination. Its extensive size and deep phenotyping further enable the development and validation of novel technologies, such as wearable sleep-monitoring technologies as well as AI applications including natural language processing approaches. *Abbreviations: AI – artificial intelligence, NLP – natural language processing, PPI – patients and public involvement*.

Since its initiation 25 years ago, the instruments, data types, and integration strategies steadily developed. Today, the sleep-wake-circadian assessments are at an exemplary level of integration with the EHCS, setting the ground for standardized data flows within the clinical routine alongside with collection of high-quality data. Complementary, clinical free-text reports from the BSWR are being analyzed using natural language processing (NLP). Two NLP-based studies have examined semantic drift within the diagnostic process for SWCD^37^ and divergence between objective and subjective measures of daytime sleepiness^38^, while recent efforts focused on enabling free-text–based ICD-11 classification of comorbidities using large language models^30^.

The BSWR provides a controlled environment for developing and validating new technologies beyond the NLP applications mentioned above. It is used to develop and test new AI-based methods, particularly for evaluating automated sleep-scoring algorithm performance^33,39,40^ and for quantifying algorithmic biases across demographic and clinical subgroups^41^. Further, the usability of wearable and nearable devices in clinical routine is evaluated^24,42,43^.

The BSWR engages in networking with national and international institutions (i.e., EUNN^44^, SNaNe^23^, KOSEK^22^) and hosts several research projects. A multimodal patient clustering over the full spectrum of sleep disorders concludes that more specific biomarkers are warranted for better patient phenotyping, and sleep co-morbidities need to be taken into consideration^45^. Also concluding with the notion that current disease markers and classification rules are insufficient, a long-term follow-up study on patients with central disorders of hypersomnolence demonstrates that Narcolepsy type 2 is an unstable diagnosis and idiopathic hypersomnia has the potential for remission^46^. Another long-term follow-up study on patients with suspicion of insufficient sleep syndrome concludes that sleep insufficiency is a condition, which should be accounted for, independent of the primary sleep disorder^47^. The availability of follow-up data further enables the “Sleep and Longevity” project, focusing on long-term outcomes and sleep microarchitecture derived digital makers^48^. Moreover, sleep dynamics derived digital markers predict future cardiovascular risk in SDB patients^49^ and reveal gender and age dependent OSA phenotypes^50^.

Next to scientific research, the BSWR supports educational initiatives and knowledge dissemination. Students enrolled to the International Master in Sleep Medicine program^25^ are introduced to the registry during their internships and can use BSWR data for academic projects. Once key milestones in data harmonization and governance are achieved, we plan to involve patients and the public in the BSWR. This will be implemented through knowledge dissemination events that share information about ongoing research projects within the BSWR and invite feedback from the public on the most relevant research directions. These initiatives will empower patients to better understand their own sleep in the context of population data and increase public awareness about sleep health.

The BSWR has limitations. Being a single-center registry, it may be subject to referral bias, with certain patient subgroups underrepresented (e.g., patients with insomnia may be referred directly to psychiatric services). The information on the prevalence of SWCD and related age- and gender aspects is provided for the purpose of the databank description but does not intend to represent epidemiological evidence and should be interpreted with caution. The BSWR is not a standardized prospective epidemiological cohort but reflects real-world clinical practice, which can result in incomplete data. Historical changes in nomenclature, including updates to ICSD classifications and AASM PSG scoring criteria, may introduce biases. Similarly, data structures have evolved over time: some parameters were only captured following the rollout of specific systems, standard procedures or questionnaires (e.g., SOMNOmedics, EPIC, FSS, BSHQ), while other features, such as systematic documentation of cataplexy symptoms, are currently de-supported. These factors need to be considered when analyzing and interpreting longitudinal or retrospective data.

## 5 Conclusion

The BSWR which is rooted in over 40 years of routine sleep medicine has transformed clinical experience into a longitudinal, multimodal resource for clinical and translational research. By integrating deep clinical phenotyping with objective sleep-wake-circadian assessments, raw physiological signals, and long-term follow-up, it provides a platform to investigate disease trajectories, identify novel biomarkers, and develop and validate data-driven technologies. As the registry continues to grow and becomes increasingly interoperable, it may help move sleep-wake-circadian medicine beyond conventional diagnostic categories toward more precise, predictive, and individualized approaches to diagnosis, prognosis, and treatment.

## Data Availability

All data produced in the present study are available upon reasonable request to the authors

## Acknowledgements

We thank Prof. Johannes Mathis and Dr. Corinne Roth for database initiation, Dr. Eduardo Villar Ortega, Uchendu Nwachukwu, Marco Pesce and Dr. Michal Bechny for data curation, and Dr. Jan Warncke, medical doctors, and study nurses for data collection. We warmly thank the patients for contributing their clinical data.

## Data Availability Statement

Data can be shared with external research groups upon reasonable request, involving a data transfer agreement describing terms of data usage and project scope.

## Ethics Statement

The study was approved by local ethical committees

## References

1. Vorster APA, van Someren EJW, Pack AI, Huber R, Schmidt MH, Bassetti CLA. Sleep Health. Clin Transl Neurosci. 2024;8(1). doi:10.3390/ctn8010008

2. Lim DC, Najafi A, Afifi L, et al. The need to promote sleep health in public health agendas across the globe. Lancet Public Heal. 2023;8(10):e820–e826. doi:10.1016/S2468-2667(23)00182-2

3. Bassetti CLA, Welter LS, Montes-Martinez M, et al. Epidemiology and Economic Burden of Sleep Disorders in Europe. Eur J Neurol. 2026;33(2):e70463. doi:10.1111/ene.70463

4. Shi L, Chen SJ, Ma MY, et al. Sleep disturbances increase the risk of dementia: A systematic review and meta-analysis. Sleep Med Rev. 2018;40:4–16. doi:10.1016/j.smrv.2017.06.010

5. Feeney SP, McCarthy JM, Petruconis CR, Tudor JC. Sleep loss is a metabolic disorder. Sci Signal. 2025;18(881):eadp9358. doi:10.1126/scisignal.adp9358

6. Lanza G, Mogavero MP, Salemi M, Ferri R. The Triad of Sleep, Immunity, and Cancer: A Mediating Perspective. Cells. 2024;13(15). doi:10.3390/cells13151246

7. Natale A, Mohanty S. Cardiovascular benefit of continuous positive airway pressure therapy in obstructive sleep apnoea: what’s new? Eur Heart J. Published online August 2025. doi:10.1093/eurheartj/ehaf460

8. Franks KH, Rowsthorn E, Nicolazzo J, et al. The treatment of sleep dysfunction to improve cognitive function: A meta-analysis of randomized controlled trials. Sleep Med. 2023;101:118–126. doi:10.1016/j.sleep.2022.10.021

9. Zhang GQ, Cui L, Mueller R, et al. The National Sleep Research Resource: towards a sleep data commons. J Am Med Inform Assoc. 2018;25(10):1351–1358. doi:10.1093/jamia/ocy064

10. Quan SF, Howard B V, Iber C, et al. The Sleep Heart Health Study: design, rationale, and methods. Sleep. 1997;20(12):1077–1085.

11. Kawamura A, Matsui K, Yoshiike T, et al. 0260 Establishment of a polysomnography databank for sleep disorders in Japan. Sleep. 2023;46(Supplement_1):A115–A116. doi:10.1093/sleep/zsad077.0260

12. Chan DECY, Bin YS, de Chazal P, et al. Sleep architecture and quality of life in comorbid OSA and depression: cross-sectional analysis of the Sydney sleep biobank. Sleep Breath. 2025;29(5):302. doi:10.1007/s11325-025-03485-y

13. Coleman RM, Roffwarg HP, Kennedy SJ, et al. Sleep-wake disorders based on a polysomnographic diagnosis. A national cooperative study. JAMA. 1982;247(7):997–1003.

14. Lee H, Li B, DeForte S, et al. A large collection of real-world pediatric sleep studies. Sci data. 2022;9(1):421. doi:10.1038/s41597-022-01545-6

15. Chen X, Wang R, Zee P, et al. Racial/Ethnic Differences in Sleep Disturbances: The Multi-Ethnic Study of Atherosclerosis (MESA). Sleep. 2015;38(6):877–888. doi:10.5665/sleep.4732

16. Young T, Palta M, Dempsey J, Skatrud J, Weber S, Badr S. The occurrence of sleep-disordered breathing among middle-aged adults. N Engl J Med. 1993;328(17):1230–1235. doi:10.1056/NEJM199304293281704

17. Saint-Maurice PF, Freeman JR, Russ D, et al. Associations between actigraphy-measured sleep duration, continuity, and timing with mortality in the UK Biobank. Sleep. 2024;47(3). doi:10.1093/sleep/zsad312

18. Marti-Soler H, Hirotsu C, Marques-Vidal P, et al. The NoSAS score for screening of sleep-disordered breathing: a derivation and validation study. Lancet Respir Med. 2016;4(9):742–748. doi:10.1016/S2213-2600(16)30075-3

19. Bonsignore MR, Hedner J. The European Sleep Apnoea Database (ESADA) ERS Clinical Research Collaboration: past, present and future. Eur Respir J. 2018;52(4). doi:10.1183/13993003.01666-2018

20. Fang R, Cheng Y, Li F, et al. The Chinese Clinical Sleep Database: An Innovative Database System Includes Large-Scale Clinical Data of Chinese Population. Nat Sci Sleep. 2024;16:305–313. doi:10.2147/NSS.S450578

21. Huang W, Wang X, Xu C, et al. Prevalence, characteristics, and respiratory arousal threshold of positional obstructive sleep apnea in China: a large scale study from Shanghai Sleep Health Study cohort. Respir Res. 2022;23(1):240. doi:10.1186/s12931-022-02141-3

22. Coordination Rare Diseases Switzerland (Kosek). https://www.kosekschweiz.ch/en

23. Bassetti CLA, Khatami R, Miano S, Werth E. The Swiss Narcolepsy Network (SNaNe). Clin Transl Neurosci. 2023;7(4):31. doi:10.3390/ctn7040031

24. Schindler KA, Nef T, Baud MO, et al. NeuroTec sitem-insel bern: closing the last mile in neurology. Clin Transl Neurosci. 2021;5(2):13. doi:10.3390/ctn5020013

25. Bassetti CLA, Sohm F, Adamantidis A, Schindler K, Barazzoni F, Manconi M. The International Master of Advanced Studies (MAS) in Sleep Medicine of the Universities of Bern and Svizzera Italiana. Clin Transl Neurosci. 2024;8(2). doi:10.3390/ctn8020019

26. Harris PA, Taylor R, Minor BL, et al. The REDCap consortium: Building an international community of software platform partners. J Biomed Inform. 2019;95:103208. doi:10.1016/j.jbi.2019.103208

27. *International Classification of Sleep Disorders (3rd Ed., Text Rev.)*. American Academy of Sleep Medicine.; 2023. https://aasm.org/clinical-resources/international-classification-sleep-disorders/

28. Valenzuela W, Balsiger F, Wiest R, Scheidegger O. Medical-Blocks-A Platform for Exploration, Management, Analysis, and Sharing of Data in Biomedical Research: System Development and Integration Results. JMIR Form Res. 2022;6(4):e32287. doi:10.2196/32287

29. Wilkinson MD, Dumontier M, Aalbersberg IJJ, et al. The FAIR Guiding Principles for scientific data management and stewardship. Sci data. 2016;3:160018. doi:10.1038/sdata.2016.18

30. Deng Y, Dennstädt F, Filchenko I, et al. Comorbidity Classification from Clinical Free-Text using Large Language Models: Application to Sleep Disorder Patients. J Med Syst. 2026;50(23). doi:10.1007/s10916-026-02343-y

31. Saver JL, Klerman EB, Buchan AM, et al. Consensus Recommendations for Standardized Data Elements, Scales, and Time Segmentations in Studies of Human Circadian/Diurnal Biology and Stroke. Stroke. 2023;54(7):1943–1949. doi:10.1161/STROKEAHA.122.041394

32. Berry RB, Brooks R, Gamaldo CE, et al. The AASM manual for the scoring of sleep and associated events. Rules, Terminol Tech Specif Darien, Illinois, Am Acad Sleep Med. 2012;176(2012):7.

33. Rossi AD, Metaldi M, Bechny M, et al. SLEEPYLAND: trust begins with fair evaluation of automatic sleep staging models. NPJ Digit Med. Published online 2025. https://arxiv.org/abs/2506.08574

34. Filchenko I, Eberhard-Moscicka AK, Picard JL, et al. Thalamic Stroke and Sleep Study: Sleep-Wake, Autonomic Regulation, and Cognition. Stroke. 2025;56(6):1528–1541. doi:10.1161/STROKEAHA.124.049156

35. Van Someren EJ, Swaab DF, Colenda CC, Cohen W, McCall W V, Rosenquist PB. Bright light therapy: improved sensitivity to its effects on rest-activity rhythms in Alzheimer patients by application of nonparametric methods. Chronobiol Int. 1999;16(4):505–518. doi:10.3109/07420529908998724

36. Lammers GJ, Bassetti CLA, Dolenc-Groselj L, et al. Diagnosis of central disorders of hypersomnolence: A reappraisal by European experts. Sleep Med Rev. 2020;52:101306. doi:10.1016/j.smrv.2020.101306

37. Deng Y, der Meer J, Tzovara A, Schmidt M, Bassetti CLA, Denecke K. Analysis of semantic drifting in diagnostic texts for sleep disorders. In: 2023 IEEE 36th International Symposium on Computer-Based Medical Systems (CBMS). 2023:726–729.

38. Deng Y, van der Meer J, Tzovara A, Schmidt MH, Bassetti CLA, Denecke K. How well did patients sleep? Analyzing sleep behavior using BERT-BiLSTM and finetuned GPT-2 Sentiment classification: a comparison study. JMIR Med INFORMATICS. 2025;13. doi:10.2196/70753

39. Fiorillo L, Monachino G, van der Meer J, et al. U-Sleep’s resilience to AASM guidelines. NPJ Digit Med. 2023;6(1):33. doi:10.1038/s41746-023-00784-0

40. Bechny M, Monachino G, Fiorillo L, et al. Bridging AI and Clinical Practice: Integrating Automated Sleep Scoring Algorithm with Uncertainty-Guided Physician Review. Nat Sci Sleep. 2024;16:555–572. doi:10.2147/NSS.S455649

41. Bechny M, Fiorillo L, van der Meer J, et al. Beyond accuracy: a framework for evaluating algorithmic bias and performance, applied to automated sleep scoring. Sci Rep. 2025;15(1):21421. doi:10.1038/s41598-025-06019-4

42. Gnarra O, Breuss A, Rossi L, et al. Transferability of a Sensing Mattress for Posture Classification from Research into Clinics. IEEE Int Conf Rehabil Robot. 2023;2023:1–6. doi:10.1109/ICORR58425.2023.10304684

43. Morand R, Gnarra O, van der Meer J, et al. iSPHYNCS: Enabling long-term actigraphy with vital signals in a comparison between Fitbit Inspire 2/HR and MotionWatch 8 for non-parametric circadian rhythm analysis. Smart Heal. 2025;38. 10.1016/j.smhl.2025.100614

44. Khatami R, Luca G, Baumann CR, et al. The European Narcolepsy Network (EU-NN) database. J Sleep Res. 2016;25(3):356–364. doi:10.1111/jsr.12374

45. Aellen FM, Van der Meer J, Dietmann A, Schmidt M, Bassetti CLA, Tzovara A. Disentangling the complex landscape of sleep-wake disorders with data-driven phenotyping: A study of the Bernese center. Eur J Neurol. 2024;31(1):e16026. doi:10.1111/ene.16026

46. Cavalli RG, Tamagni AEM, van der Meer J, et al. Narcolepsy type 2 is an unstable diagnosis and idiopathic hypersomnia has the potential for remission - long-term and cross-sectional observations from the Bern Sleep−Wake Registry. Sleep. Published online August 2025. doi:10.1093/sleep/zsaf242

47. Wenz ES, Fregolente L, Kälin V, van der Meer J, Schmidt MH, Bassetti CLA. Insufficient sleep syndrome: A single center (Bern) experience over 20 years in diagnosis and management. Submitt to Sleep Med. Published online 2026.

48. Filchenko I, van der Meer J, Bernasconi CA, Schmidt MH, Bassetti CLA. Sleep and longevity: Insights from sleep macroarchitecture and nocturnal heart rate variability. Eur J Neurol. 2025;32. doi:10.1111/ene.70189

49. Bechny M, Kishi A, Tomita Y, et al. Sleep-Stage Dynamics Predict Current Sleep-Disordered Breathing and Future Cardiovascular Risk. medRxiv. Published online 2025. doi:10.1101/2025.07.31.25332545

50. Bechny M, Kishi A, Fiorillo L, et al. Novel digital markers of sleep dynamics: causal inference approach revealing age and gender phenotypes in obstructive sleep apnea. Sci Rep. 2025;15(1):12016. doi:10.1038/s41598-025-97172-3

