## Supplementary material for "The Bern Sleep-Wake Registry: A 25-year multimodal resource for data-driven sleep medicine"

[1]Sleep-Wake Epilepsy Center, Department of Neurology, Inselspital, University Hospital Bern, University of Bern, Bern, Switzerland.
[2]Department of Pulmonary Medicine, Allergology and Clinical Immunology, Inselspital, Bern University Hospital, University of Bern, Bern 3010, Switzerland
[3]Department of Neurology, Kufstein Hospital, Kufstein, Austria
[4]Clinics for Sleep Medicine Luzern, Zurzach Care, Lucerne, Switzerland
[5]Graduate School for Cellular and Biomedical Sciences, University of Bern, Switzerland
[6]Institute of Computer Science, University of Bern, Bern, Switzerland
[7]Institute of Digital Technologies for Personalized Healthcare (MeDiTech), University of Applied Sciences and Arts of Southern Switzerland (SUPSI), Lugano, 6962, Switzerland
[8]Neurology Department, Neurocenter of Southern Switzerland, Ente Ospedaliero Cantonale, 6903, Switzerland
[9]Sleep Research Institute, Madrid, Spain
[10]Institute for Diagnostic and Interventional Neuroradiology, Inselspital, Bern University Hospital, University of Bern, Bern, Switzerland
[11]University Hospital of Psychiatry, University of Bern, Bern, Switzerland
[12]Università della Svizzera italiana, Lugano, Switzerland
[§]First author, corresponding author
*Last author

### Supplementary Material

**TABLE S1 |** Demographics, medical history, and living status.

| Parameter | Data type | Data source | Available from |
|---|---|---|---|
| Unique patient-level record_id | Numeric | REDCap | 2021 |
| Birth year | Numeric | IDSC | 2000 |
| Birth month | Numeric | IDSC | 2000 |
| Sex | Categorical | IDSC | 2000 |
| Complete history of diagnoses | Text | IDSC | 2010 |
| Medication history | Text | IDSC | 2010 |
| Smoking | Text | IDSC | 2010 |
| Alcohol consumption | Text | IDSC | 2010 |
| Availability of laboratory samples | Boolean | IDSC | 2010 |
| General consent status | Categorical | IDSC | 2000 |

Listed are parameters, data types, data source and year from which data are available. Data are obtained from the electronic health care systems (EHCS) via the Insel data science center (IDSC).

**TABLE S2 |** Questionnaires.

| Questionnaire | Abbreviation | Introduction date | Condition |
|---|---|---|---|
| Epworth sleepiness scale | ESS | 2000 | - |
| Fatigue severity scale | FSS | 2012 | - |
| Swiss narcolepsy scale | SNS | 2004 | IND |
| World Health Organization-Five Well-Being Index | WHO-5 | 2023 | - |
| Insomnia Severity Index | ISI | 2023 - 2025 | - |
| Bern Sleep-Health Questionnaire | BSHQ | 2023 | - |
| Patient Health Questionnaire-9 | PHQ-9 | 2023 | SH |
| Generalized Anxiety Disorder Scale-7 | GAD-7 | 2023 | SH |
| International RLS Scale | iRLS | 2023 | IND |
| Composite Morningness Questionnaire | CMQ | 2023 | SH |

Listed are questionnaires, abbreviations, introduction date and condition (to minimize the burden of patients, some questionnaires are only presented if indicated based on scores from other questionnaires (Condition = IND) and some questionnaires are only presented to patients consulting via the Swiss Sleep House (Condition = SH).

**TABLE S3 |** Overnight PSG parameters.

| Parameter | Data type | Introduction date |
|---|---|---|
| Start date | Date | 2000 |
| Breathing control | Categorical | 2000 |
| Subjective sleep quality (PSG night) | Categorical | 2000 |
| Subjective sleep quality (night before test) | Categorical | 2022 |
| Subjective sleep duration (PSG night) | Categorical | 2008 |
| Subjective sleep latency (PSG night) | Categorical | 2022 |
| PSG Report | Text | 2000 |
| Lights off | Time | 2000 |
| Lights on | Time | 2000 |
| Total sleep time (minutes) | Numeric | 2000 |
| Sleep latency (minutes) | Numeric | 2000 |
| REM latency (minutes from sleep onset) | Numeric | 2000 |
| Stage transition Index (n/h) | Numeric | 2000 |
| Sleep efficiency (%) | Numeric | 2021 |
| Wake (%TIB) | Numeric | 2021 |
| REM (%TST) | Numeric | 2021 |
| Non-REM 1 (%TST) | Numeric | 2000 |
| Non-REM 2 (%TST) | Numeric | 2000 |
| Non-REM 3 (%TST) | Numeric | 2000 |
| REM (%TST) | Numeric | 2000 |
| Body position supine (%TST) | Numeric | 2021 |
| AHI (n/h) | Numeric | 2000 |
| AHI REM (n/h) | Numeric | 2021 |
| AHI NREM (n/h) | Numeric | 2021 |
| AHI supine (n/h) | Numeric | 2021 |
| AHI non-supine (n/h) | Numeric | 2021 |
| Apnea index (n/h) | Numeric | 2000 |
| Hypopnea index (n/h) | Numeric | 2000 |
| Central apnea index (n/h) | Numeric | 2021 |
| Obstructive apnea index (n/h) | Numeric | 2021 |
| Mixed apnea index (n/h) | Numeric | 2021 |
| Mean SpO2 (%) | Numeric | 2000 |
| Minimal SpO2 (%) | Numeric | 2000 |
| Duration saturation < 90% (minutes) | Numeric | 2000 |
| Duration saturation < 88% (minutes) | Numeric | 2021 |
| Duration saturation < 80% (minutes) | Numeric | 2000 |
| ODI (n/h) | Numeric | 2000 |
| Mean tCO2 (mmHg) | Numeric | 2005 |
| Maximal tCO2 (mmHg) | Numeric | 2005 |
| Mean heart frequency (bpm) | Numeric | 2000 |
| Arousal index (n/h) | Numeric | 2021 |
| PLMS Index (n/h) | Numeric | 2005 |

Listed are parameters, data types and year from which data are available.

*Abbreviations:* AHI - Apnea Hypopnea Index; SpO2 – Peripheral oxygen saturation; ODI – Oxygen desaturation index; tCO2 – Transcutaneous carbon dioxide; PLMS – Periodic limb movements during sleep.

**TABLE S4 |** Objective parameters of daytime sleepiness.

| Parameter | Data type | Repetitions | Introduction date |
|---|---|---|---|
| **MSLT** | | | |
| MSLT Start date | Date | 1 | 2000 |
| Number of MSLT tests | Categorical | 1 | 2000 |
| Indication for MSLT test | Categorical | 1 | 2000 |
| MSLT Report | Text | 1 | 2000 |
| MSLT Start time | Time | 4 or 5 | 2000 |
| MSLT Sleep latency to any sleep stage (from lights-off) | Numeric | 4 or 5 | 2000 |
| MSLT REM latency (from sleep onset) | Numeric | 4 or 5 | 2000 |
| MSLT Latency to sleep stage N1 (minutes from lights-off) | Numeric | 4 or 5 | 2021 |
| MSLT Latency to sleep stage N2 (minutes from lights-off) | Numeric | 4 or 5 | 2021 |
| MSLT Latency to sleep stage N3 (minutes from lights-off) | Numeric | 4 or 5 | 2021 |
| MSLT Sleep duration (minutes) | Numeric | 4 or 5 | 2000 |
| MSLT KSS before | Numeric | 4 or 5 | 2004 |
| MSLT KSS after | Numeric | 4 or 5 | 2004 |
| MSLT Mean sleep latency (minutes) | Numeric | 1 | 2000 |
| MSLT Number of epochs with sleep (n) | Numeric | 1 | 2000 |
| MSLT Mean REM latency (minutes) | Numeric | 1 | 2000 |
| MSLT Number of epochs with REM sleep (n) | Numeric | 1 | 2000 |
| **MWT** | | | |
| MWT Start date | Date | 1 | 2002 |
| Number of MWT tests (n) | Categorical | 1 | 2002 |
| Indication for MWT test | Categorical | 1 | 2002 |
| MWT Report | Text | 1 | 2002 |
| MWT Start time | Time | 4 or 5 | 2002 |
| MWT Sleep latency to any sleep stage (minutes from lights-off) | Numeric | 4 or 5 | 2002 |
| MWT Occurrence of REM sleep | Boolean | 4 or 5 | 2002 |
| MWT KSS before | Numeric | 4 or 5 | 2004 |
| MWT KSS after | Numeric | 4 or 5 | 2004 |
| MWT Mean sleep latency (minutes) | Numeric | 1 | 2002 |
| MWT Number of epochs with sleep (n) | Numeric | 1 | 2002 |
| MWT Number of epochs with REM sleep (n) | Numeric | 1 | 2002 |
| **Vigilance** | | | |
| PVT mean RT (ms) | Numeric | 1 | 2005 |
| PVT lapses (n) | Numeric | 1 | 2005 |
| SART (total errors) | Numeric | 1 | 2018 |
| Stear clear test (% errors) | Numeric | 1 | 2005 |
| Pupillography (variation) | Numeric | 1 | 2010 |

Listed are parameters, data types, number of repetitions within a test battery and year from which data are available. For repeated measures, parameters for each repetition are available. Results from multiple sleep latency tests (MSLT), maintenance of wakefulness tests (MWT) and vigilance tests are reported. Patients indicate a Karolinska Sleepiness Score (KSS) immediately before and after the MSLT / MWT round. For the psychomotor vigilance test (PVT) mean reaction times (RT) and lapses are reported. For the Sustained Attention to Response Task (SART), total errors are reported. For the Stear clear driving ability test, % errors are reported, for pupillography, the mean variation is reported.

**TABLE S5 |** Wrist actigraphy parameters.

| Parameter | Data type | Introduction date |
|---|---|---|
| Device type | Categorical | 2000 |
| Actigraphy start date | Date | 2000 |
| Actigraphy duration (days) | Numeric | 2000 |
| Inactivity Index (%) | Numeric | 2000 |
| Average lights-off time | Time | 2024 |
| Minimal lights-off time | Time | 2024 |
| Maximal lights-off time | Time | 2024 |
| Average time falling asleep (minutes) | Time | 2024 |
| Average lights-on time | Time | 2024 |
| Minimal lights-on time | Time | 2024 |
| Maximal lights-on time | Time | 2024 |
| Average time in bed (minutes) | Numeric | 2024 |
| Minimal time in bed (minutes) | Numeric | 2024 |
| Maximal time in bed (minutes) | Numeric | 2024 |
| Mean sleep efficiency (%TIB) | Numeric | 2024 |
| Mean wake bout time (seconds) | Numeric | 2024 |
| Total number of naps (n) | Numeric | 2024 |
| Relative amplitude (NPCR) | Numeric | 2024 |
| Inter-daily stability (NPCR) | Numeric | 2024 |
| L5 start (NPCR) | Time | 2024 |
| M10 start (NPCR) | Time | 2024 |

Listed are parameters, data types, and year from which data are available.

*Abbreviations: TIB – time in bed; NPCR* - non-parametric circadian rhythm analysis; L5 - least active 5 hours; M10 – most active 10 hours

**TABLE S6 |** General information from sleep-wake-circadian assessments.

| Parameter | Data type | Data source | Introduction date |
|---|---|---|---|
| Visit start | Date | Derived from assessment dates | 2016 |
| Visit end | Date | Derived from assessment dates | 2016 |
| Age | Numeric | IDSC | 2000 |
| Height (cm) | Numeric | SOMNOmedics / IDSC | 2000 |
| Weight (kg) | Numeric | SOMNOmedics / IDSC | 2000 |
| BMI (kg/m$^2$) | Numeric | Calculated | |
| Waist circumfence (cm) | Numeric | SOMNOmedics | 2000 |
| Hip circumfence (cm) | Numeric | SOMNOmedics | 2000 |
| Medication at electrophysiological testing | Text | SOMNOmedics | 2000 |
| Clinical history | Text | SOMNOmedics | 2021 |
| Referring cause | Text | IDSC | 2000 |
| Clinical summary | Text | IDSC | 2021 |
| Clinical procedure | Text | IDSC | 2021 |

Listed are parameters, data types, data source and year from which data are available. Main data sources are the electronic health care systems (EHCS) via the Insel data science center (IDSC) and SOMNOmedics.

**TABLE S7 |** Sleep-wake-circadian disorder classification.

| Parameter | Data type | Scope | Repetitions |
|---|---|---|---|
| Event-level diagnosis | Categorical | Event-level | 5 |
| Event-level diagnosis certainty | Categorical | Event-level | 5 |
| Global diagnosis | Categorical | Patient-level | 5 |
| Global diagnosis certainty | Categorical | Patient-level | 5 |

SWCD were assigned according to ICSD-3, which remains challenging for several reasons. Because SWCD may evolve over time, diagnoses are tracked at baseline as well as in follow-up visits (indicated as event-level diagnoses / scope). Because sleep comorbidities are frequent, up to 5 SWCD diagnoses (indicated as repetitions) may be given per visit (primary, secondary, tertiary, quaternary and quinary diagnoses). Diagnosis certainty may be depicted as definite, probable or possible. We consolidate diagnoses on patient level by prioritizing diagnoses from the most recent event (indicated as global diagnosis / patient-level scope). To establish the presence of sleep disorders, we consolidate information from the comorbid global SWCD diagnoses. If diagnoses are only suspected (certainty level “possible”), they are not considered for analysis.
